# The Efficacy of Early Magnesium Sulphate Catharsis in Potential Severe Acute Pancreatitis: A Clinical Cohort Study

**DOI:** 10.64898/2026.04.29.26352029

**Authors:** Guojian Yin, Yuxin Liu, Heng Yang, Jiaming Wu, Shuqi Qin, Si Dai, Li Cui, Jing Zhou, Jiujing Huang, Fengjie Ji, Wan Pu, Zhendan Wang, Hanqin Chen, Kewei Hu

## Abstract

**Background and Aims:** Acute pancreatitis (AP) can lead to systemic inflammatory response (SIRS), paralytic intestinal obstruction, and in severe cases, intra-abdominal hypertension (IAH), organ failure (OF), and even death. Early magnesium sulphate (MS) catharsis treatment can relieve paralytic intestinal obstruction and IAH, thus reducing the incidence of severe acute pancreatitis (SAP) as well as mortality.

**Methods:** 850 patients with AP at high risk of systemic complications (potential SAP (p-SAP)) were recruited. These p-SAP patients were categorized into two groups based on the treatment they received: the routine group (RT group, standard treatment for AP) and the MS group (early MS catharsis added to standard treatment). The final cohort had an allocation ratio of 2.5:1 (RT : MS). The primary composite endpoints were clinically finally diagnosed SAP (d-SAP) and mortality. The intensive care unit (ICU) admission, OF, inflammatory factors, length of hospital stay, and hospitalization costs, etc. were also compared and analyzed.

**Results:** It demonstrated that early MS catharsis treatment was highly effective in preventing patients with p-SAP of various aetiologies from deteriorating to d-SAP and reducing their mortality significantly. In addition, it significantly reduced the incidence of OFs, pancreatic necrosis, ICU admissions, CRRT utilisation, and the cost and length of stay of hospitalisation for these patients.

**Conclusions:** It showed that early MS catharsis had a significant therapeutic effect on p-SAP patients, which would profoundly contribute to the clinical management of AP.

## 1. INTRODUCTION

Acute pancreatitis (AP) is currently one of the most common causes for acute gastrointestinal hospitalisation (1). The incidence of AP ranges from 10 to 100 cases per 100,000 populations in different countries (2), which is still increasing at a rate of 3.07% per year (3). 10%–20% of AP patients will deteriorate to severe acute pancreatitis (SAP) (4, 5), with irreversible organ failure (OF), uncontrollable infections, and mortality up to 30%–40% (6). These will significantly increase healthcare costs, imposing additional burdens on people and the healthcare system (7).

SAP patients often experience systemic inflammatory response (SIRS), paralytic intestinal obstruction, intra-abdominal hypertension (IAH), multiple organ failure (MOF), and in severe cases, death (8). De Waele *et al*. found that 78% of SAP patients were likely to suffer from IAH (9). And patients with IAH had a higher incidence of OFs (9), including ARDS (95% *vs*. 33%, *P* = 0.004), heart failure (91% *vs*. 17%, *P* = 0.001), and renal failure (86% *vs*. 17%, *P* = 0.004) than those without IAH. Another study found that SAP patients with IAH also had a significantly higher mortality rate than patients without IAH (36% *vs*. 0%, *P* < 0.01) (10). Intestinal mucosal damage, intestinal and retroperitoneal oedema, and intestinal bacterial translocation may also occur during this process (11). All of these conditions can seriously impede the application of early enteral nutrition and affect its effectiveness (12).

So far, there have been no effective methods in previous reports and guidelines to address these issues. A hypothesis was first proposed in this study that early magnesium sulphate (MS) catharsis treatment may alleviate paralytic intestinal obstruction, avoid the occurrence of IAH, alleviate intestinal and peri-intestinal oedema, and ultimately reduce the severity of AP. MS is a hypertonic laxative that has previously been used for bowel preparation during colonoscopy (13). This clinical cohort study explored the therapeutic effect of early MS catharsis on AP and has achieved good results.

## 2. PARTICIPANTS AND METHODS

### 2.1. DIAGNOSIS, SEVERITY, AND ETIOLOGICAL CLASSIFICATION OF AP

Diagnostic criteria and severity classification of AP is based on the revised Atlanta Classification of Acute Pancreatitis (14).

Etiological classification of AP: biliary (15), hypertriglyceridemic (16), and others (3).

### 2.2. CRITERIA FOR INCLUSION AND EXCLUSION

Inclusion Criteria: All adults with AP at high risk of systemic complications (defined as potential SAP (p-SAP). Candidates need to meet one of the following three criteria within the first 3 days after onset: a. Acute Physiology and Chronic Health Evaluation II (APACHE II) scores (17) **≥** 8; b. the Imrie Score or Modified Glasgow Score (18) ≥ 3; c. serum C-reactive protein (CRP) > 150 mg/L (19).

Exclusion Criteria: a. Patients admitted to the hospital more than 48 hours after the onset of AP; b. Comorbidity with other chronic or severe diseases (including chronic heart failure, chronic renal failure, and other serious infections, etc.); c. AP due to endoscopic retrograde cholangiopancreatography or with chronic pancreatitis; d. withdrawal from the study; e. Age less than 18 years or more than 75 years.

### 2.3. STUDY DESIGN AND PROCEDURES

Patients attending the Second Affiliated Hospital of Soochow University from 1 January 2018 to 31 December 2023 who meet the inclusion and exclusion criteria and agreed to participate in the study.

Patients in the RT group received standard treatment according to the AP guidelines (8, 14) and did not receive any catharsis treatment.

Patients in the MS group received MS catharsis treatment immediately, once admitted to the hospital, on the basis of standard treatment. These patients took 50 ml of MS solution orally or through a nasogastric tube every 2 hours (25% concentration, 2.5 g/10 ml/shot, Yangzhou Zhongbao Medicine Co., China) until the faecal evacuation, relief of abdominal pain, and paralytic intestinal obstruction. Patients can use up to 6 times per day (2.5 g * 5 times * 6 times = 75 g). Afterwards, the patients can start oral enteral nutrition (12), and keep the patients’ bowel movements 2–3 times a day by continuing to take small doses of MS orally.

### 2.4. STUDY ENDPOINTS

The primary endpoints are the composite of mortality and clinically finally diagnosed SAP (d-SAP).

Secondary endpoints: organ failure (OF) (20); pancreatic necrosis score (20, 21); intensive care unit (ICU) admission; Continuous Renal Replacement Therapy (CRRT) needed; C-reactive protein (CRP), procalcitonin (PCT), amylase, and lipase levels etc., which were measured on days 1, 2, 3, and 6 after onset of AP; the length and cost of hospitalisation (see protocol for specific definitions).

### 2.5. DATA COLLECTION AND END POINT ASSESSMENT

A review committee composed of five gastroenterologists in the AP field evaluated the occurrence of primary and secondary endpoints for each patient to conduct the final analysis. Several experienced radiologists independently interpreted all CT/MRI examination results. Detailed clinical characteristics were collected, including age, gender, duration of hospitalisation, cost, laboratory data, comorbidities (hypertension, diabetes mellitus, fatty liver), and serious complications (ARDS, heart failure, renal failure, and MOF), etc.

Considering that the death of patients during the acute phase of AP may affect the comparison of some indicators, such as duration and cost of hospitalisation. In order to ensure the accuracy of the statistical results, these patients were excluded when comparing hospitalisation time and expenses.

### 2.6. PATIENT SAFETY

An independent data and safety monitoring committee assessed the progress of the study and examined the safety endpoints of 242 patients in the MS group, without any adverse event.

### 2.7. STATISTICAL ANALYSIS

The statistical analyses were performed using SPSS version 29.0.2.0 (IBM, Armonk, NY, USA). All continuous variables were shown as mean ± standard deviation (SD). Continuous-type variable data conforming to normal distribution were tested using Student’s t-test, and for those not obeying normal distribution, the Mann-Whitney U test was used as a non-parametric test. Categorical variables, all expressed as numbers and percentages, were tested by the Pearson χ^2^ test and Fisher’s exact χ^2^ test (both two-tailed tests). The binary indicators were calculated by binary logistic regression analysis and expressed as odds ratios (OR), 95% confidence intervals (CI), and P-values. P values less than 0.05 were regarded as statistically significant.

## 3. RESULTS

### 3.1. CHARACTERISTICS OF THE PATIENTS

A total of 862 p-SAP patients participated in this study (Figure S1), including 608 patients in the RT group and 242 patients in the MS group (Table 1).

**Table 1.**
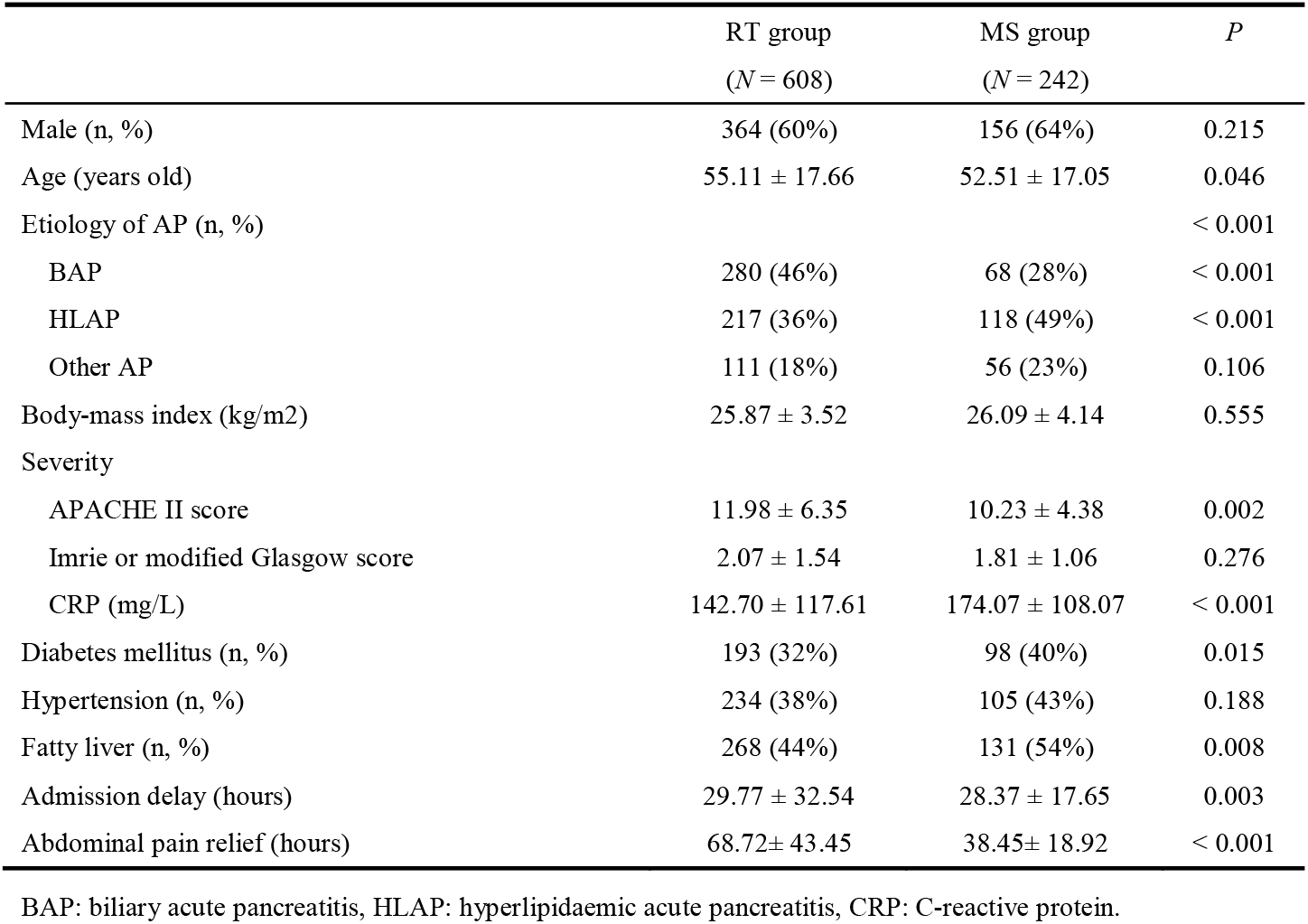
The baseline characteristics of the RT group and MS group.

The MS group had more HLAP patients (118, 49%), compared with the RT group (217, 36%). This might be related to a lower mean age (22-24), a higher CRP level (25, 26), and more patients with diabetes mellitus and fatty liver in the MS group when compared to the MS group (40-42). The average admission time of patients in the MS group is slightly earlier than that in the RT group (28.37 ± 17.65 *vs*. 29.77 ± 32.54).

### 3.2. OUTCOMES OF p-SAP PATIENTS

#### 3.2.1. Primary End Points

The primary composite endpoint (mortality and d-SAP) occurred in 101 patients (17%) in the RT group, compared to 11 patients (5%) in the MS group (absolute risk difference [ARD]: 12%, *P* < 0.001; OR: 0.239, 95% CI: 0.126–0.454, *P’* < 0.001) (Table 2). The incidence of d-SAP in the RT group was significantly higher than that in the MS group (17% *vs*. 5%, *P* < 0.001). 12 (2%) patients eventually died in the RT group, but 0 (0%) in the MS group (*P* = 0.024).

**Table 2.**
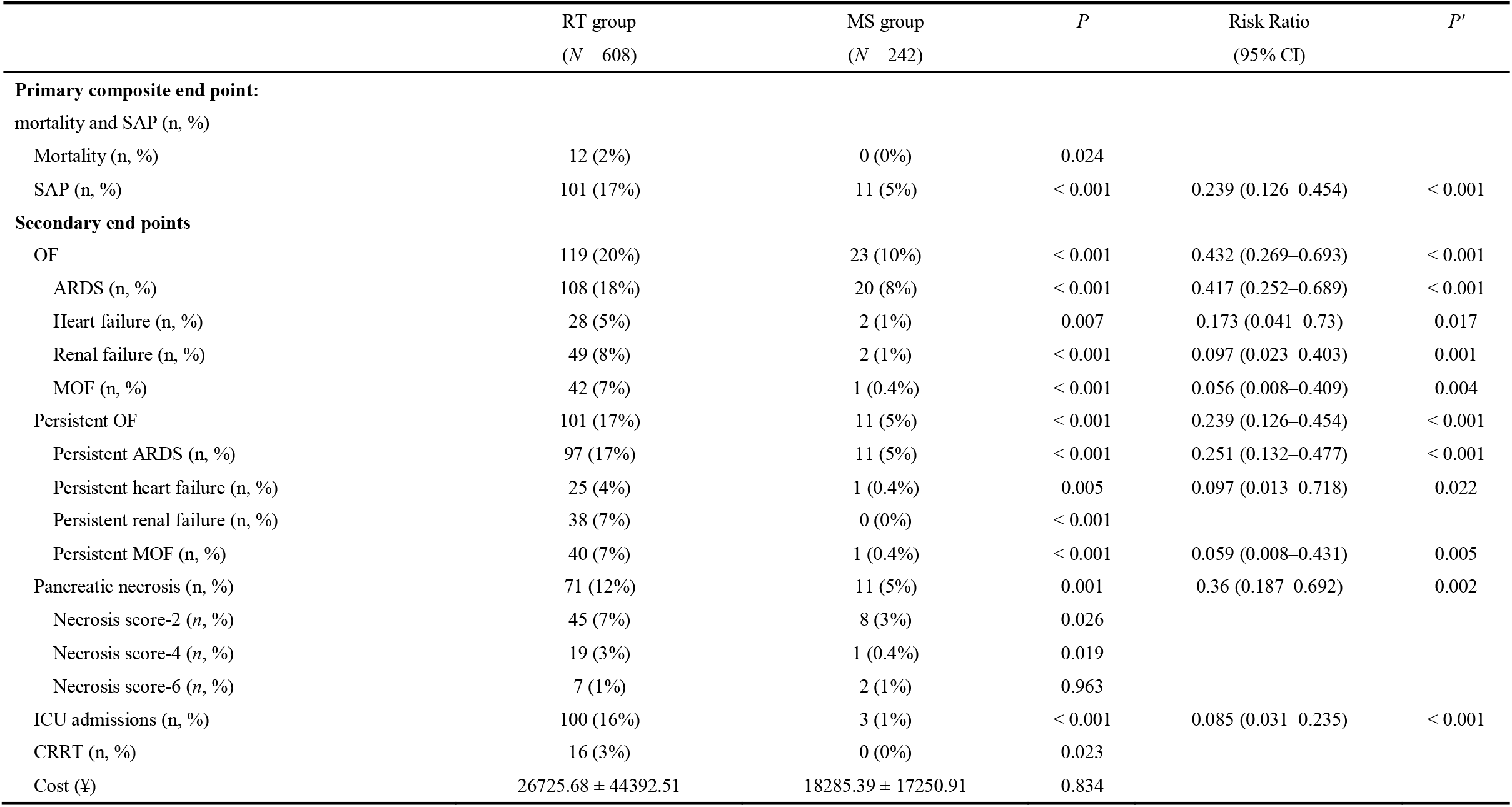
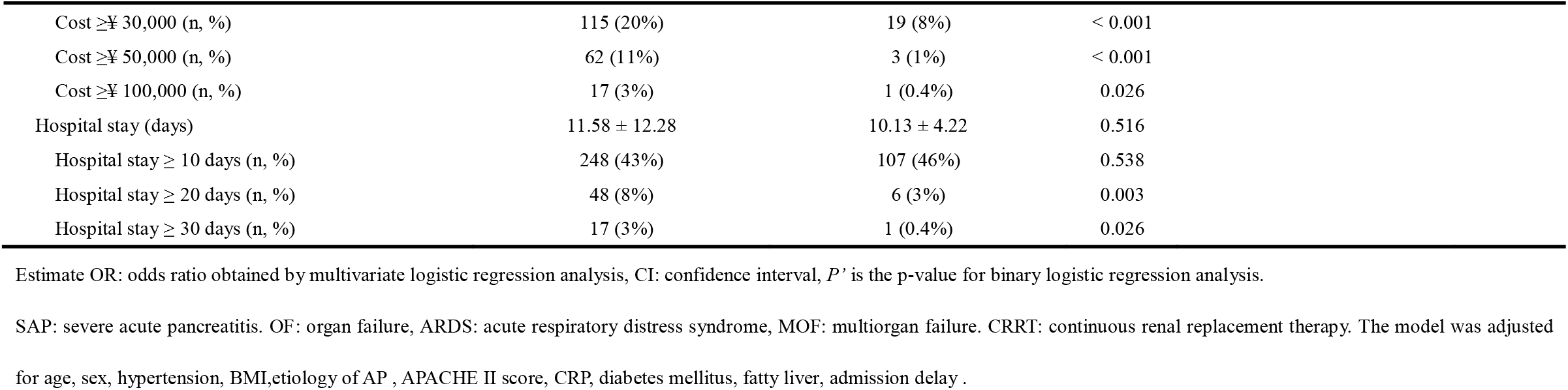
Comparison of clinical data between two treatment regimens for patients with p-SAP.

|  | RT group<br>(N = 608) | MS group<br>(N = 242) | P | Risk Ratio<br>(95% CI) | P' |
| --- | --- | --- | --- | --- | --- |
| **Primary composite end point:** |  |  |  |  |  |
| mortality and SAP (n, %) |  |  |  |  |  |
| Mortality (n, %) | 12 (2%) | 0 (0%) | 0.024 |  |  |
| SAP (n, %) | 101 (17%) | 11 (5%) | < 0.001 | 0.239 (0.126–0.454) | < 0.001 |
| **Secondary end points** |  |  |  |  |  |
| OF | 119 (20%) | 23 (10%) | < 0.001 | 0.432 (0.269–0.693) | < 0.001 |
| ARDS (n, %) | 108 (18%) | 20 (8%) | < 0.001 | 0.417 (0.252–0.689) | < 0.001 |
| Heart failure (n, %) | 28 (5%) | 2 (1%) | 0.007 | 0.173 (0.041–0.73) | 0.017 |
| Renal failure (n, %) | 49 (8%) | 2 (1%) | < 0.001 | 0.097 (0.023–0.403) | 0.001 |
| MOF (n, %) | 42 (7%) | 1 (0.4%) | < 0.001 | 0.056 (0.008–0.409) | 0.004 |
| Persistent OF | 101 (17%) | 11 (5%) | < 0.001 | 0.239 (0.126–0.454) | < 0.001 |
| Persistent ARDS (n, %) | 97 (17%) | 11 (5%) | < 0.001 | 0.251 (0.132–0.477) | < 0.001 |
| Persistent heart failure (n, %) | 25 (4%) | 1 (0.4%) | 0.005 | 0.097 (0.013–0.718) | 0.022 |
| Persistent renal failure (n, %) | 38 (7%) | 0 (0%) | < 0.001 |  |  |
| Persistent MOF (n, %) | 40 (7%) | 1 (0.4%) | < 0.001 | 0.059 (0.008–0.431) | 0.005 |
| Pancreatic necrosis (n, %) | 71 (12%) | 11 (5%) | 0.001 | 0.36 (0.187–0.692) | 0.002 |
| Necrosis score-2 (n, %) | 45 (7%) | 8 (3%) | 0.026 |  |  |
| Necrosis score-4 (n, %) | 19 (3%) | 1 (0.4%) | 0.019 |  |  |
| Necrosis score-6 (n, %) | 7 (1%) | 2 (1%) | 0.963 |  |  |
| ICU admissions (n, %) | 100 (16%) | 3 (1%) | < 0.001 | 0.085 (0.031–0.235) | < 0.001 |
| CRRT (n, %) | 16 (3%) | 0 (0%) | 0.023 |  |  |
| Cost (¥) | 26725.68 ± 44392.51 | 18285.39 ± 17250.91 | 0.834 |  |  |

|  |  |  |  |
| --- | --- | --- | --- |
| Cost ≥¥ 30,000 (n, %) | 115 (20%) | 19 (8%) | < 0.001 |
| Cost ≥¥ 50,000 (n, %) | 62 (11%) | 3 (1%) | < 0.001 |
| Cost ≥¥ 100,000 (n, %) | 17 (3%) | 1 (0.4%) | 0.026 |
| Hospital stay (days) | 11.58 ± 12.28 | 10.13 ± 4.22 | 0.516 |
| Hospital stay ≥ 10 days (n, %) | 248 (43%) | 107 (46%) | 0.538 |
| Hospital stay ≥ 20 days (n, %) | 48 (8%) | 6 (3%) | 0.003 |
| Hospital stay ≥ 30 days (n, %) | 17 (3%) | 1 (0.4%) | 0.026 |

Estimate OR: odds ratio obtained by multivariate logistic regression analysis, CI: confidence interval, *P*’ is the p-value for binary logistic regression analysis.
SAP: severe acute pancreatitis. OF: organ failure, ARDS: acute respiratory distress syndrome, MOF: multiorgan failure. CRRT: continuous renal replacement therapy. The model was adjusted
for age, sex, hypertension, BMI,etiology of AP , APACHE II score, CRP, diabetes mellitus, fatty liver, admission delay .

Figure 2 C1 showed the difference of severity of AP between the two groups. It was evident that MS catharsis significantly reduced the odds of developing SAP in p-SAP patients (OR: 0.239, 95% CI: 0.126–0.454; *P’* < 0.001).

**Figure 2.**
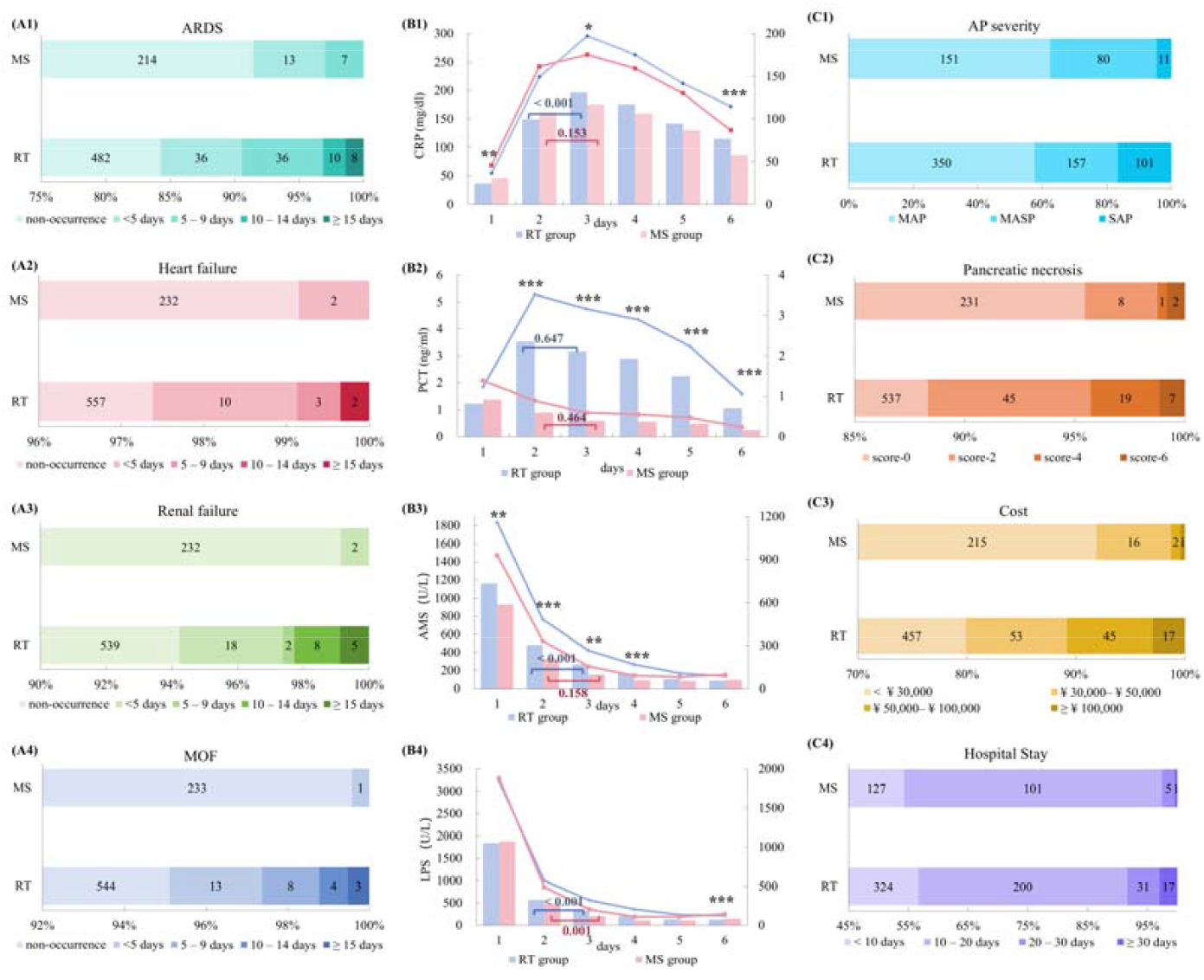
The baseline characteristics of p-SAP patients between RT group and MS group. (A1–A4) Number of patients with ARDS, heart failure, renal failure, MOF, and duration. The darker the colour block, the longer the duration of the complication. (B1–B4) Levels of CRP (C-reactive protein), PCT (Procalcitonin), AMS (amylase), and LPS (lipase) in R T and MS groups. Comparison of RT and MS groups: * *P* < 0.05, ** *P* < 0.01, *** *P* < 0.001. (C1–C4) AP severity, pancreatic necrosis, cost, and hospital stay in the RT group and MS groups.

#### 3.2.2. Secondary End Points

##### 3.2.2.1. Organ failure

Compared with the RT group, p-SAP patients in the MS group had a significantly lower incidence of all types of OFs, especially renal failure (ARD, 7%; OR: 0.097) and MOF (ARD, 6.6%; OR: 0.056). And the risk of persistent OF in the MS group was 0.239 times lower than that in the RT group (ARD, 12%; 95% CI: 0.126–0.454; *P’* < 0.001). For persistent MOF, MS catharsis remained effective (ARD, 6.6%; OR: 0.059; *P’* = 0.005).

The duration of OFs in both groups was presented in Figure 2 A1–A4. More patients in the RT group had prolonged courses of OFs.

##### 3.2.2.2. Pancreatic necrosis

Pancreatic necrosis was observed in 5% of the p-SAP patients in the MS group, compared to 12% in the RT group. It was significant that the treatment of MS catharsis was effective (OR: 0.36, 95% CI: 0.187–0.692; *P’* = 0.002), detailed in Fig. 2 C2.

##### 3.2.2.3. ICU admissions & CRRT use

After early MS catharsis, the proportion of p-SAP patients needed to be admitted to ICU decreased from 16% to 1%, with a direct reduction of risk to 0.085-fold (95% CI: 0.031–0.235; *P’* < 0.001). The p-SAP patients in the MS group did not require CRRT (0%), compared to 3% in the RT group. Only two patients in the MS group developed renal failure, and both recovered within 48 h.

##### 3.2.2.4. Cost & Hospital stay

Patients in the MS group have shorter hospital stays and lower costs. Figure 2 C4 showed that there were more patients in the RT group hospitalised for more than 30 days (3% *vs*. 0.4%). 8% of p-SAP patients in the RT group had costs between ¥ 50,000–¥ 100,000, and 3% of patients had costs ≥ ¥ 100,000, compared to 1% and 0.3% in the MS group, respectively (Figure 2 C3).

##### 3.2.2.5. Inflammatory indicators

In the MS group, both CRP and PCT decreased more rapidly (Table S1 and Fig. 2. B1–B2). There was a significant difference of CRP between days 2 and 3 in the RT group (*P* < 0.001), but no difference in the MS group (*P* = 0.153), indicating that inflammation was controlled more rapidly after MS catharsis. The amylase and lipase in the MS group also recovered to the normal range faster (Fig. 2.B3–B4).

### 3.3. THE CLINICAL OUTCOMES IN d-SAP PATIENTS AFTER MS TREATMEN

MS treatment is also effective for those patients with finally diagnosed SAP (d-SAP). Patients with d-SAP in the MS group had a more rapid decline in inflammatory markers, less incidence and duration of OFs, and fewer hospitalisations and costs, when compared to those d-SAP patients in the RT group (Fig. 3).

**Figure 3.**
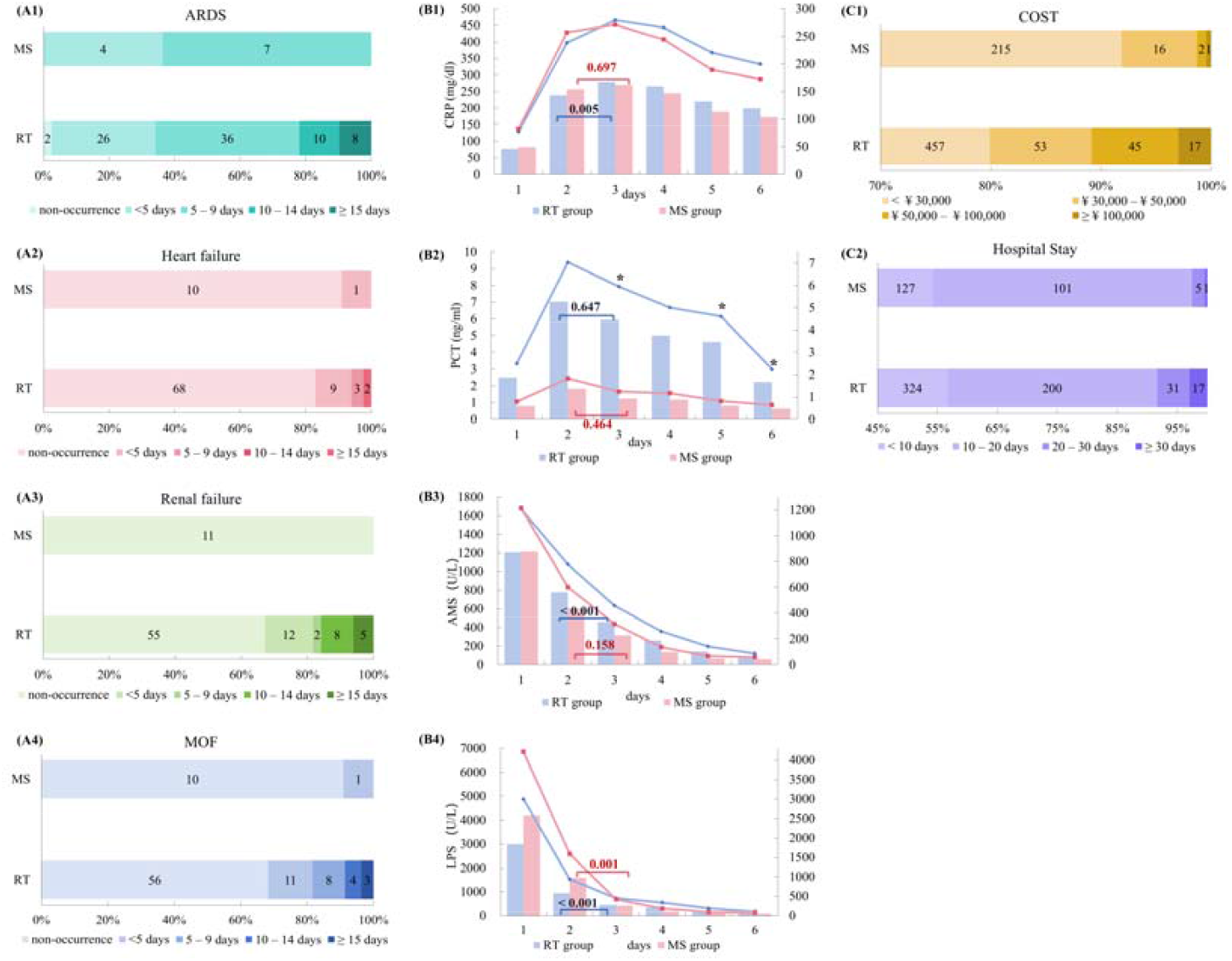
The clinical outcomes of d-SAP patients after MS administration in the two groups. (A1–A4) Numbers of patients with ARDS, heart failure, renal failure, MOF, and duration of organ failure. The darker the colour block, the longer the duration of the organ failure. (B1–B4) Levels of CRP (C-reactive protein), PCT (Procalcitonin), AMS (amylase), and LPS (lipase) in R T and MS groups. Comparison of RT and MS groups: * *P* < 0.05. (C1–C2) Costs and hospital stays in the RT group and MS groups.

### 3.4. THE CLINICAL OUTCOMES FOR p-SAP PATIENTS WITH DIFFERENT AETIOLOGIES

#### 3.4.1. BAP

68 p-SAP patients with BAP were in the MS group (233.30 ± 107.12 ml of MS used on average), and 280 patients were in the RT group. In the RT group, 9 patients died (3%), and 16% of patients were finally diagnosed with d-SAP, compared to no deaths and 4% of d-SAP in the MS group (Table 3). There were fewer patients with OF in the MS group (7% *vs*. 19%; *P1* = 0.019), especially with ARDS and renal failure. The durations of OFs in both groups were shown in Figure S2. There were fewer patients who needed ICU admission in the MS group (6% *vs*. 15%) and CRRT utilization (0% *vs*. 2%).

**Table 3:**
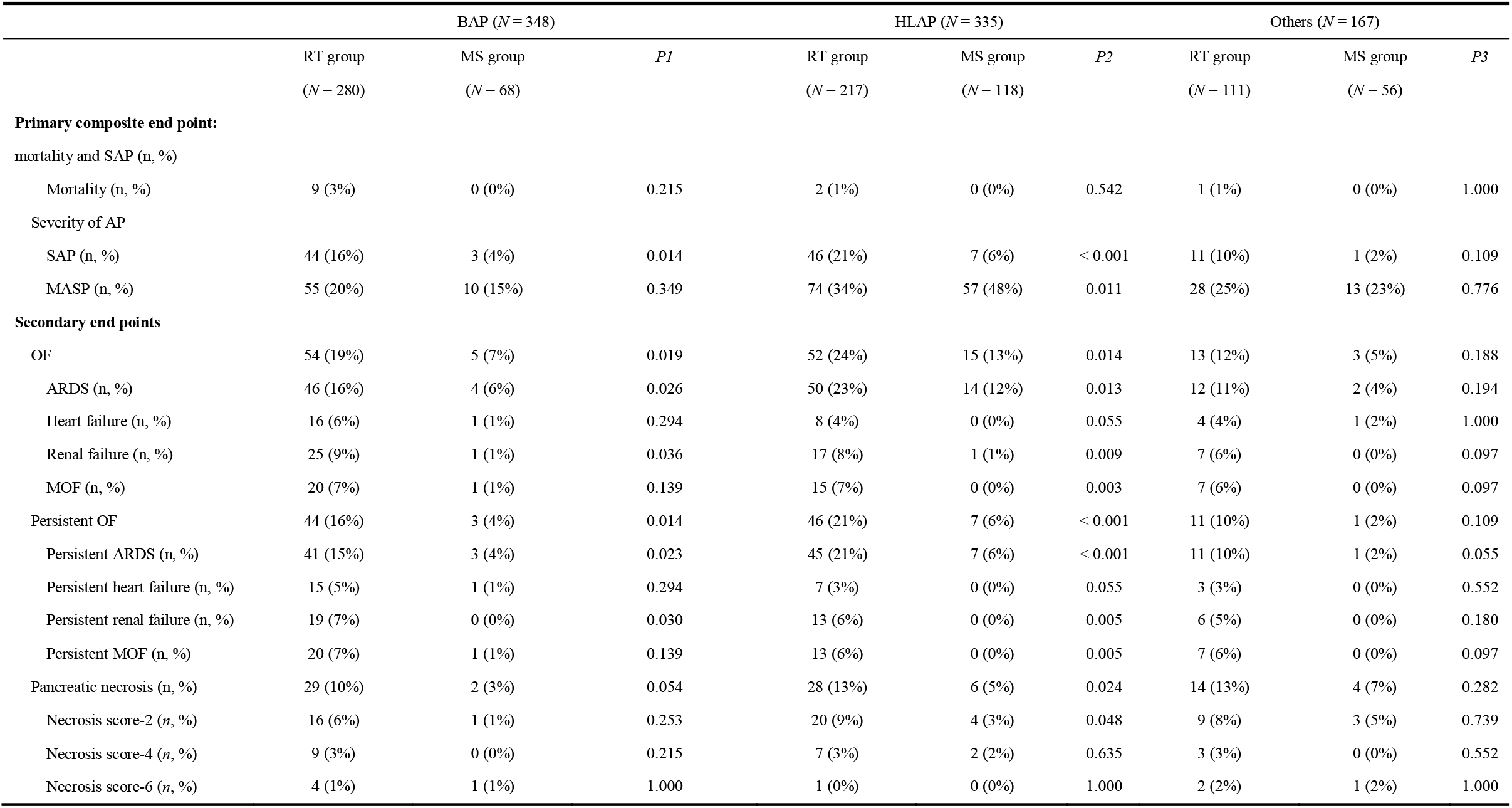
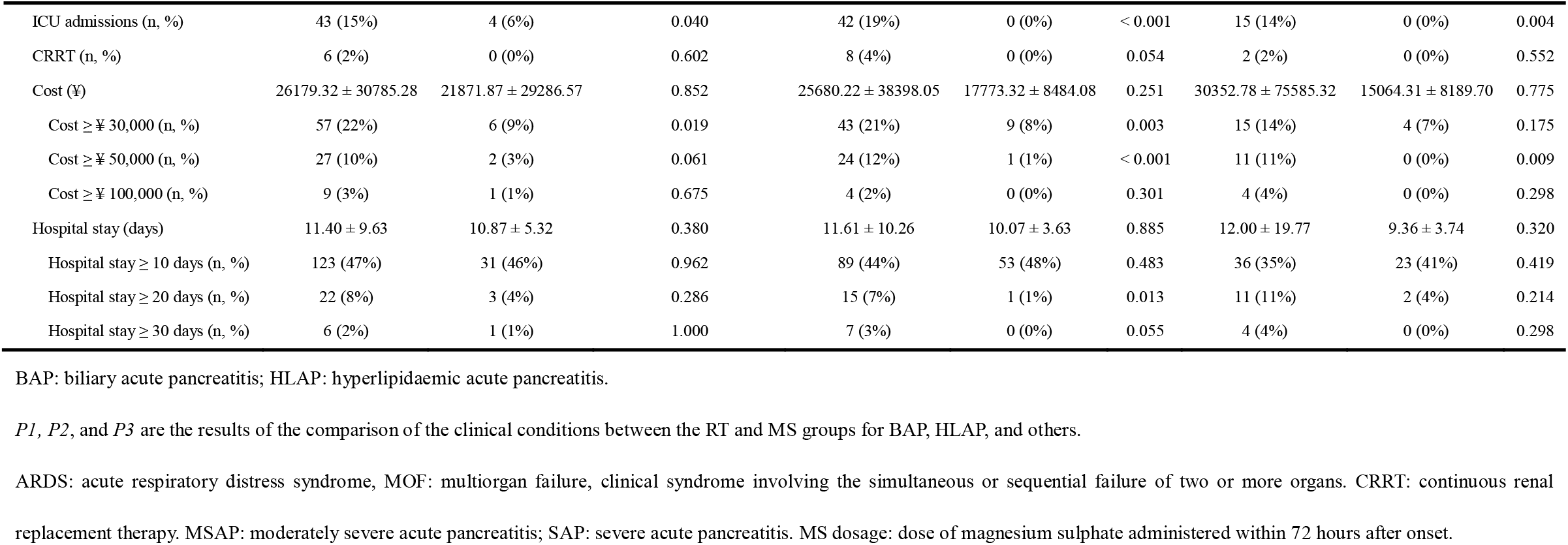
Differences between the two groups in patients with AP of different aetiologies.

#### 3.4.2. HLAP

In the RT group, 21% of patients developed d-SAP, and two died, while in the MS group (195.90 ± 89.06 ml of MS used on average), only 6% developed d-SAP (*P2* = 0.011), and all 118 patients survived. The incidence of persistent ARDS (ARD, 15%; *P2* <0.001) and persistent renal failure (ARD, 6%; *P2* =0.005) were also significantly reduced in the MS group. The duration of OFs in the MS group was significantly shorter (Fig. S2. B1–4). No one in the MS group developed MOF, while 7% of patients in the RT group did.

#### 3.4.3. Other etiologies

10% of patients in the RT group deteriorated to d-SAP, compared to 2% in the MS group, with an average oral dose of 208.90 ± 89.75 ml of MS. No one in the MS group was admitted to ICU (ARD, 14%; *P3* = 0.004).

## 4. DISCUSSION

The cohort study proposed that early MS catharsis treatment had a significant therapeutic effect on p-SAP patients.

MS is a common clinical medication, once commonly used for intestinal cleansing before colonoscopy. In this study patients with p-SAP were given 50 ml of MS solution (solution concentration: 25%) orally every 2 hours immediately after admission until faecal evacuation, abdominal pain, and paralytic intestinal obstruction resolved. The mean MS dosage of the patients in the MS group during the first 3 days after onset was 209.30 ± 95.40 ml (Q1 = 150 ml, Q2 = 200 ml, Q3 = 250 ml). The patients in the MS group showed significant relief of abdominal pain at 38.45 ± 18.92 h (Q1 = 25 h, Q2 = 30 h, Q3 = 50 h), compared to the patients in the RT group, who required 68.72 ± 43.45 h (Q1 = 32 h, Q2 = 65 h, Q3 = 97 h).

It was found that the early MS catharsis was a definitely effective treatment for p-SAP patients. Both the mortality rate and the incidence of d-SAP in the MS group were significantly lower than those in the RT group (Table 2). The mortality rate was 0% in the MS group compared to 2% (12) in the RT group (*P* = 0.024). Only 5% of p-SAP patients in the MS group deteriorated to d-SAP, compared to 17% in the RT group, with a risk reduction of 0.239-fold (ARD, 12%; 95% CI: 0.126–0.454; *P’* < 0.001). The patients in the MS group also had a lower risk of developing OF (Table 2) and a shorter duration of OFs (Fig. 2.A1–4). 10% of patients in the MS group and 20% in the RT group suffered from OF (ARD, 10%, *P* < 0.001), suggesting that MS lowered the risk of OF to 0.432-fold (95% CI: 0.269–0.693; *P* < 0.001). In addition, patients in the MS group showed a faster decline in inflammatory indicators and lower peak levels (Figure 2 B1–4 and Table S1). It also confirmed that MS laxatives are equally effective in treating p-SAP patients with different etiologies (Table 3).

In the United States, the average hospitalisation cost for AP exceeds $30,000 (27, 28), while in China it is approximately up to ¥60,000 (29). In this study, it can be seen that MS treatment significantly reduced the number of patients who spent over ¥30,000 (Fig. 2 C3 and Fig. 3 C1), with an average cost of ¥18,285.39 ± 17,250.91 for all those p-SAP patients, rather than those patients with AP in the broad sense. The ICU admission rate and CRRT utilisation rate in the MS group were significantly reduced, with ARD rates of 15% and 3%, respectively. From a socio-economic perspective, patients with MS catharsis had better clinical outcomes, which greatly reduced their costs and time of hospitalisation.

In this study, early MS catharsis was strongly recommended to treat AP, and it was found that MS had many advantages. Firstly, moderate to severe pancreatitis usually suffers from paralytic intestinal obstruction, which reduces blood perfusion, damages intestinal mucosal integrity and barrier function, and promotes the progression and aggravation of systemic inflammation (30). Although paralytic intestinal obstruction plays a crucial role in the progression of SAP, there are still no effective treatments in previous reports and guidelines to address the vicious cycle of paralytic intestinal obstruction and the following IAH. Early MS can effectively alleviate paralytic intestinal obstruction in the acute phase of AP, allowing for early oral enteral nutrition and promoting early recovery of intestinal function (12). Secondly, MS catharsis can cleanse the intestines, thereby reducing the risk of intestinal bacterial translocation. Fritz et al. (39) found that decontamination of the small intestine in rats significantly reduced the rate of pancreatic reinfection (P<0.005), while decontamination of the colon had almost no effect, indicating that the main source of bacterial translocation and infectious pancreatic necrosis is the small intestine rather than the colon. Finally, MS has high safety (31), and we have not observed any renal toxicity or electrolyte imbalance in the clinical use (32).

In short, patients with early MS catharsis treatment had faster recovery, lower cost, fewer ICU admissions, and extremely lower d-SAP incidence rate and mortality, which could greatly and significantly improve the prognosis of patients.

## Abbreviations List

AP: acute pancreatitis
BAP: biliary acute pancreatitis
HLAPAL: hyperlipidemic acute pancreatitis
MAP: mild acute pancreatitis
MSAP: moderately severe acute pancreatitis
SAP: severe acute pancreatitis
p-SAP: potential SAP
d-SAP: finally diagnosed SAP
ALT: alanine aminotransferase
ULN: upper limit of normal
OF: organ failure
MOF: multiple organ failure
HF: heart failure
RF: renal failure
CRP: C-reactive protein
PCT: procalcitonin
RT group: routine group
MS group: magnesium sulphate group
OR: odds ratio
CI: confidence interval
ARD: absolute risk reduction.

## Data availability statement

The datasets generated and/or analyzed during this study are available upon request from the corresponding author.

## Author contributions

Yuxin Liu: formal analysis, investigation, data curation, statistical analysis, writing-original draft; Heng Yang: data curation, writing-original draft. Yuxin Liu and Heng Yang contribut ed equally to this article. Shuqi Qin: investigation, data curation; Si Dai: investigation, dat a curation; Jing Zhou: investigation, data curation; Jiujing Huang: data curation, Fengjie Ji: data curation, Wan Pu: data curation; Zhendan Wang: data curation, Hanqin Chen: patient diagnosis and treatment, observation of therapeutic effect, follow-up of patients, Kewei H u: patient diagnosis and treatment, observation of therapeutic effect, follow-up of patients, Guojian Yin*: Proposal, design, and implementation of research, writing-review & editing, Funding.

## Acknowledgements

We thank the Second Affiliated Hospital of Soochow University for providing case data for this study.

## Ethical approval

The study was approved by the Ethics Committee of the Second Affiliated Hospital of Soochow University (JD-LK-2022-064-02) and was conducted in accordance with GCP principles.

## Participant informed consent statement

All subjects have been informed and signed informed consent to participate this study.

## Consent for publication

All subjects and authors agreed to the final manuscript and its submission to the journal.

---

**Figure S1.**
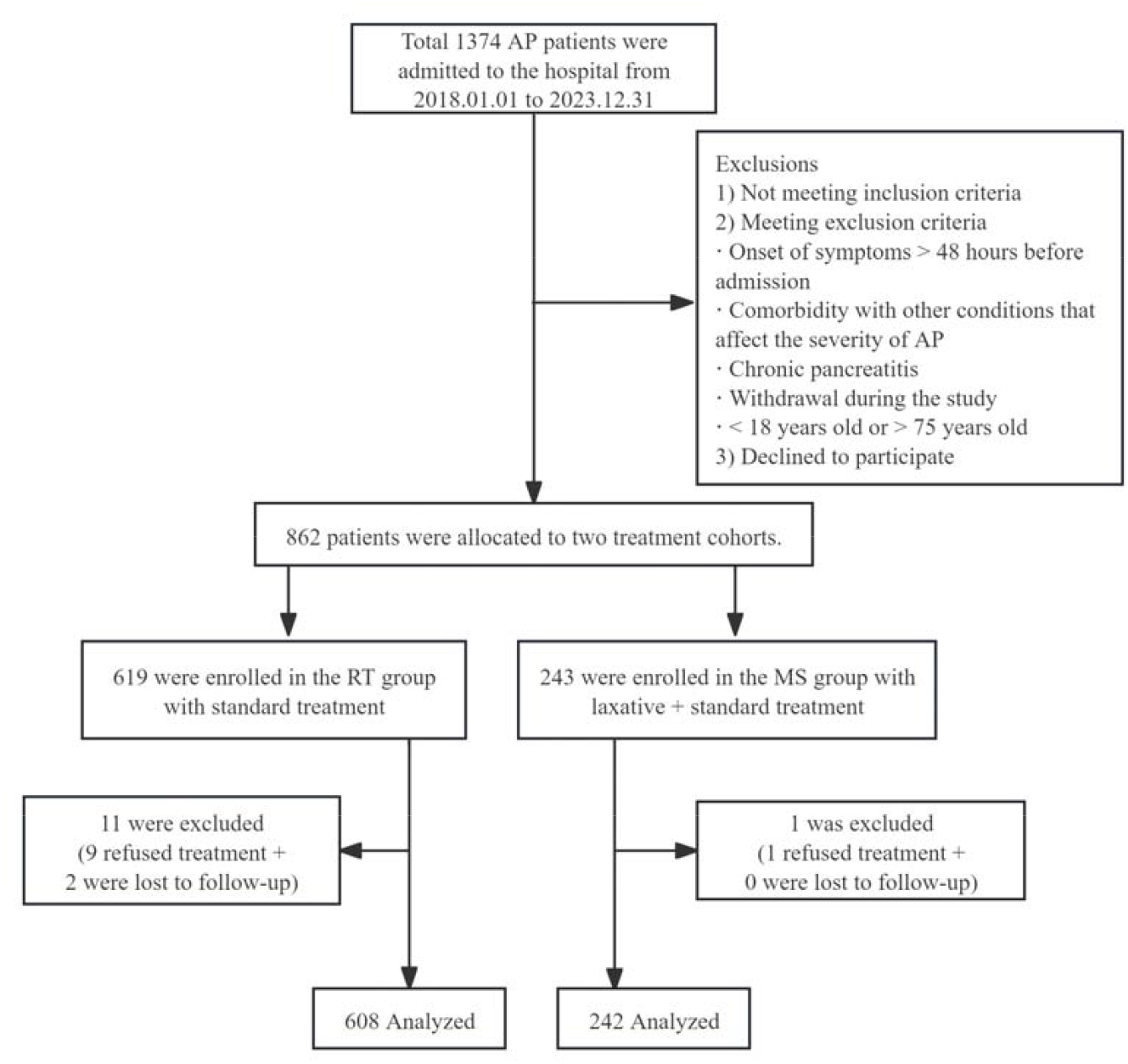
Study flaw chart.

**Table S1.**
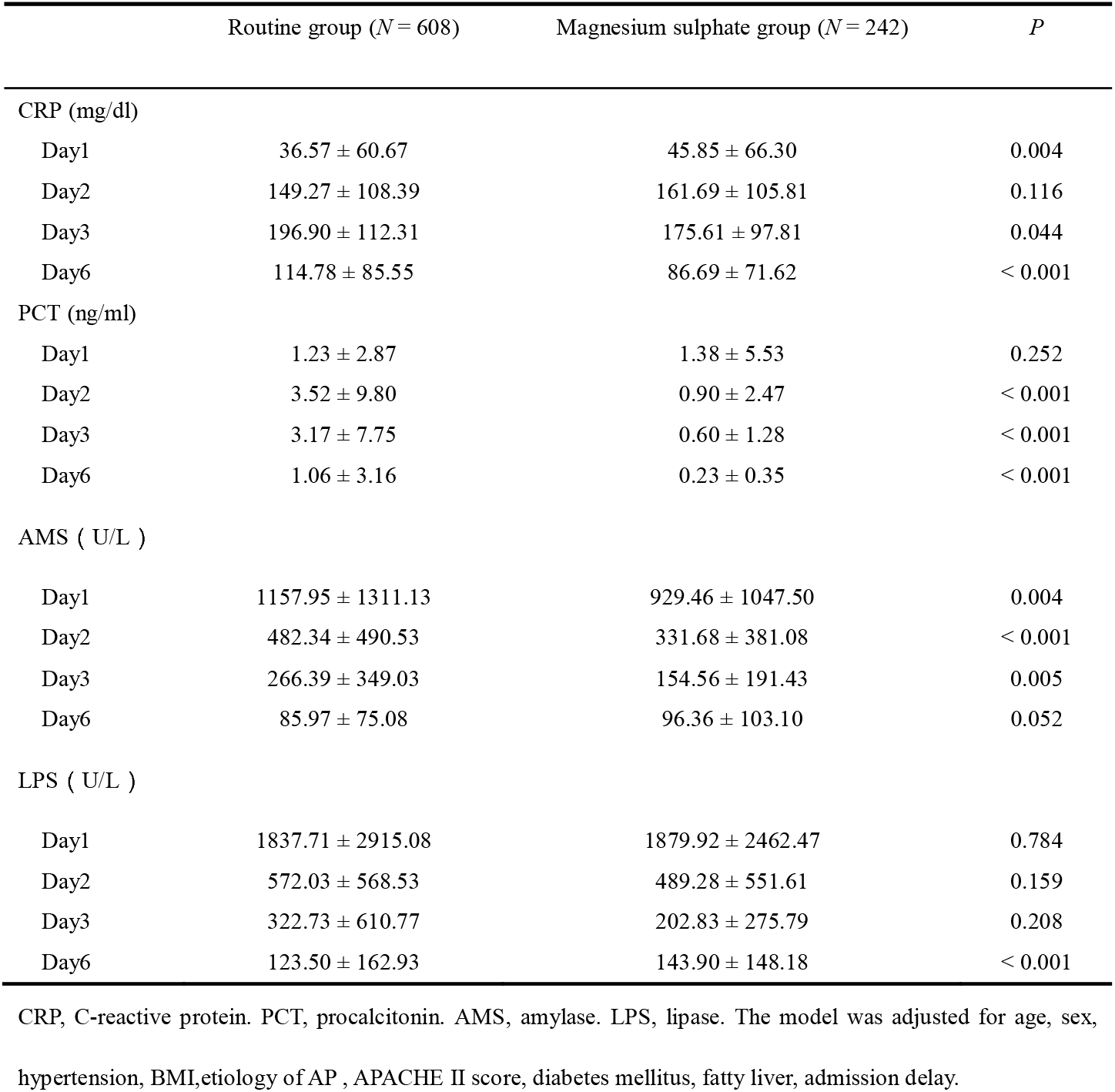
Comparison of inflammation indicators between two groups for p-SAP patients.

|  | Routine group (*N* = 608) | Magnesium sulphate group (*N* = 242) | *P* |
| --- | --- | --- | --- |
| CRP (mg/dl) |  |  |  |
| Day1 | 36.57 ± 60.67 | 45.85 ± 66.30 | 0.004 |
| Day2 | 149.27 ± 108.39 | 161.69 ± 105.81 | 0.116 |
| Day3 | 196.90 ± 112.31 | 175.61 ± 97.81 | 0.044 |
| Day6 | 114.78 ± 85.55 | 86.69 ± 71.62 | < 0.001 |
| PCT (ng/ml) |  |  |  |
| Day1 | 1.23 ± 2.87 | 1.38 ± 5.53 | 0.252 |
| Day2 | 3.52 ± 9.80 | 0.90 ± 2.47 | < 0.001 |
| Day3 | 3.17 ± 7.75 | 0.60 ± 1.28 | < 0.001 |
| Day6 | 1.06 ± 3.16 | 0.23 ± 0.35 | < 0.001 |
| AMS ( U/L ) |  |  |  |
| Day1 | 1157.95 ± 1311.13 | 929.46 ± 1047.50 | 0.004 |
| Day2 | 482.34 ± 490.53 | 331.68 ± 381.08 | < 0.001 |
| Day3 | 266.39 ± 349.03 | 154.56 ± 191.43 | 0.005 |
| Day6 | 85.97 ± 75.08 | 96.36 ± 103.10 | 0.052 |
| LPS ( U/L ) |  |  |  |
| Day1 | 1837.71 ± 2915.08 | 1879.92 ± 2462.47 | 0.784 |
| Day2 | 572.03 ± 568.53 | 489.28 ± 551.61 | 0.159 |
| Day3 | 322.73 ± 610.77 | 202.83 ± 275.79 | 0.208 |
| Day6 | 123.50 ± 162.93 | 143.90 ± 148.18 | < 0.001 |

CRP, C-reactive protein. PCT, procalcitonin. AMS, amylase. LPS, lipase. The model was adjusted for age, sex,
hypertension, BMI,etiology of AP , APACHE II score, diabetes mellitus, fatty liver, admission delay.

